# Skeletal muscle properties in long COVID and ME/CFS differ from those induced by bed rest

**DOI:** 10.1101/2025.05.02.25326885

**Authors:** Braeden T. Charlton, Anouk Slaghekke, Brent Appelman, Moritz Eggelbusch, Jelle Y. Huijts, Wendy Noort, Paul W. Hendrickse, Frank W. Bloemers, Jelle J. Posthuma, Paul van Amstel, Richie P. Goulding, Hans Degens, Richard T. Jaspers, Michèle van Vugt, Rob C.I. Wüst

## Abstract

Patients with long COVID and myalgic encephalomyelitis/chronic fatigue syndrome (ME/CFS) suffer from a reduced exercise capacity, skeletal muscle abnormalities and post-exertional malaise (PEM), where symptoms worsen with cognitive or physical exertion. PEM often results in avoidance of physical activity, resulting in a lower aerobic fitness, which may contribute to skeletal muscle abnormalities. Here, we compared whole-body exercise responses and skeletal muscle adaptations after strict 60-day bed rest in healthy people with those in patients with long COVID and ME/CFS, and healthy age- and sex-matched controls. Bed rest altered the respiratory and cardiovascular responses to (sub)maximal exercise, while patients exhibited respiratory alterations only at submaximal exercise. Bed rest caused muscle atrophy, and the reduced oxidative phosphorylation related to reductions in maximal oxygen uptake. Patients with long COVID and ME/CFS did not have muscle atrophy, but had less capillaries and a more glycolytic fibers, none of which were associated with maximal oxygen uptake. While the whole-body aerobic capacity is similar following bed rest compared to patients, the skeletal muscle characteristics differed, suggesting that physical inactivity alone does not explain the lower exercise capacity in long COVID and ME/CFS.

## Introduction

Although most acute SARS CoV2 infections resolve within days to weeks, some symptoms persist or worsen in a subset of patients^1–3^. The continuation or development of symptoms after 12 weeks is termed long COVID, and patients typically experience debilitating fatigue, brain fog, postural orthostatic tachycardia, myalgia, and post-exertional malaise (PEM)^3^. PEM, experienced by ∼90% of patients with long COVID^4^, is the worsening of symptoms following physical or psychological exertion. While the pathophysiology underlying long COVID remains unclear, recent evidence suggests a close resemblance to myalgic encephalomyelitis/chronic fatigue syndrome (ME/CFS)^5^, another disease characterized by reduced exercise capacity, brain fog and PEM. Importantly, PEM often results in avoidance of physical activity, which may contribute to the skeletal muscle abnormalities often observed in both conditions^6–12^. Exercise intensity and duration largely dictates the magnitude of whole-body exercise^13^ and skeletal muscle adaptations^14^; however due to PEM, long COVID and ME/CFS patients may not achieve such intensities or durations frequently enough to induce such adaptations, resulting in deconditioning^15–17^.

Deconditioning can range from mild step reductions to strict bed rest, with more severe models leading to more rapid reductions in aerobic capacity and skeletal muscle function^18–21^. These declines are often associated with muscle atrophy, capillary rarefaction, as well as reduced mitochondrial content and mitochondrial function^22–25^, but also reduced cardiac output^23,26^ and maximal ventilatory capacity^27,28^. However, whether these alterations cohere with those in patients with long COVID and ME/CFS has not been directly investigated. Patients with long COVID and ME/CFS exhibit lower aerobic exercise capacity and mitochondrial respiration than healthy individuals^6,7,9,10^, reduced capillary densities^9^, and some indications of muscle atrophy^7^, which has often been attributed to deconditioning^15,16^ However, whether deconditioning alone accounts for these changes remains unclear.

The current study aimed to compare whole body exercise responses and skeletal muscle adaptations in patients with long COVID and ME/CFS to those observed following strict 60-day bed rest. We hypothesized that while whole body aerobic capacity would be similarly lower between patients and following bed rest, adaptations in skeletal muscle markers for oxygen supply and utilization would diverge. Specifically, we expected patients to display no muscle atrophy, and reduced capillary-to-fiber ratios and capillary densities. In contrast, we expected bed rest to result in severe muscle atrophy, reduced capillary-to-fiber ratios, and increased capillary density. Further, we expected that muscle size and tissue markers for oxygen supply and utilization rates are associated to whole-body exercise capacity in healthy participants undergoing bed rest, but that such association is lost in patients. To test these hypotheses, we analysed aerobic exercise responses and vastus lateralis skeletal muscle biopsies of patients with ME/CFS and long COVID, and compared these to age- and sex-matched healthy controls, as well as a cohort of 24 healthy individuals who completed 60-days of strict bed rest.

## Results

### Participant Characteristics

Table 1 provides the participant characteristics of the two cohorts, Figure 1A provides a graphical overview of the cohorts and study design. The healthy controls and patients with long COVID and ME/CFS were all vaccinated against SARS CoV2 at the time of measurements, while the bed rest study was conducted before the COVID pandemic. Patients with long COVID developed symptoms prior to vaccination, and met the Canadian Consensus Criteria for ME/CFS. Patients with ME/CFS were already diagnosed with ME/CFS before the COVID pandemic, and were therefore ill for a significantly longer time relative to long COVID patients. Both patient groups displayed mild symptoms, as is implied by their willingness and ability to undergo exercise testing. Patients with long COVID and ME/CFS displayed large interindividual differences in daily step count (range: 733-8609 steps/day; Table 1).

**Figure 1:**
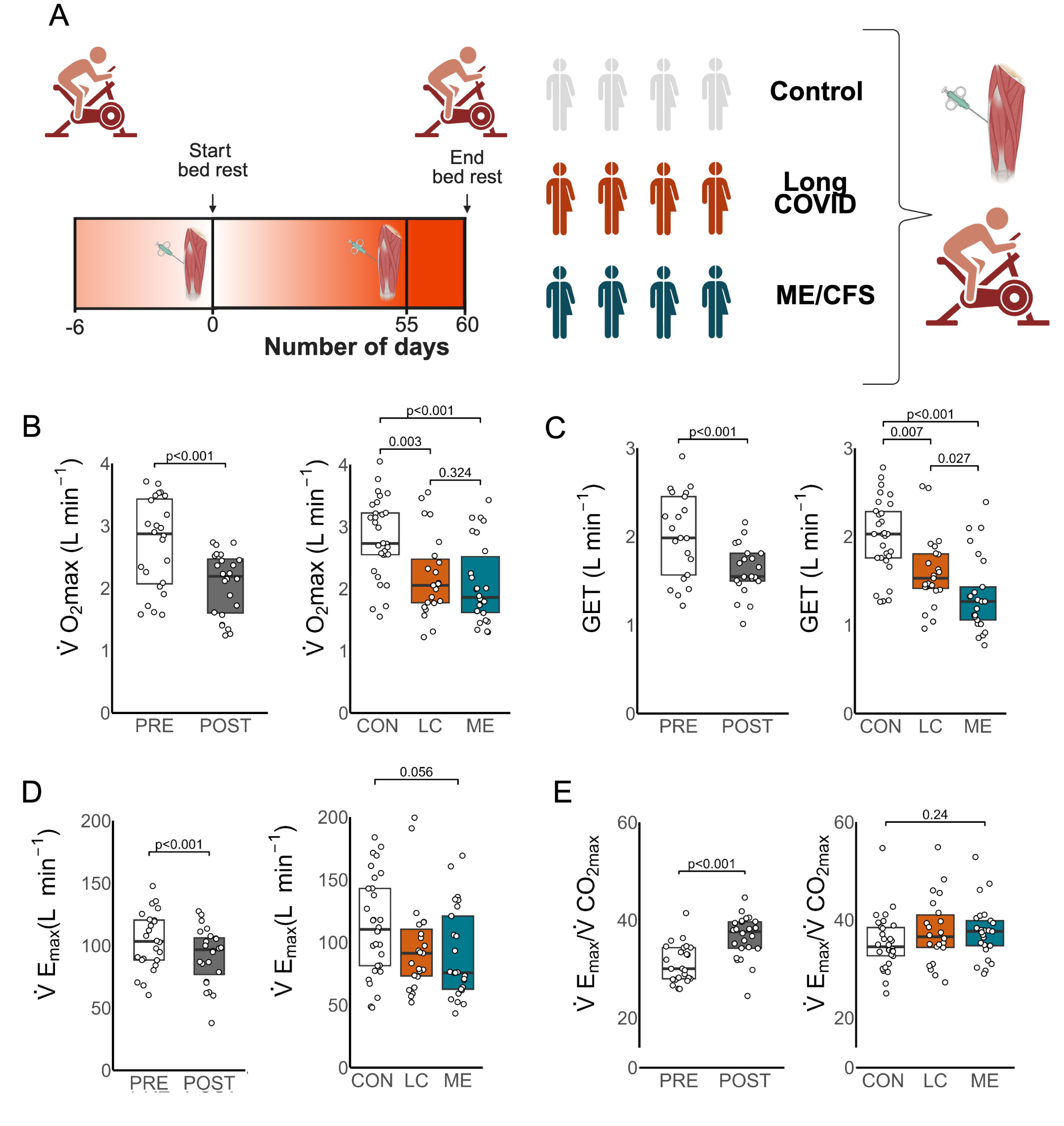
**A**: Study designs for the bed rest and the ME/CFS and long COVID cohorts. Bed rest decreased 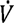*0*_2*max*_ (**B**), gas exchange threshold (GET), and 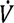*E_max_* (**C**), while 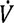*E_max_* /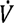*CO*_2*max*_ increased (**D**). Patients with long COVID and ME/CFS had lower 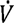*O*_2*max*_ (**B**) and GET (**C**) compared to controls and 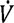*E* tended to be lower (**C**), however 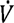*E_max_*/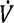*CO*_2*max*_ was not significantly different than controls (**D**). Only 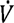*O*_2*max*_ data was non-normally distributed following Box-Cox transformations. Comparisons between pre- and post-bed rest were assessed using paired t-tests for parametric data and Mann–Whitney *U* test for non-parametric data. Comparisons between patients with long COVID and patients with ME/CFS and healthy controls were assessed using analysis of variance, with Tukey HSD post-hoc testing for parametric data or Kruskal–Wallis *H* test, with pairwise Wilcoxon tests with Benjamini–Hochberg correction post-hoc for non-parametric data.

**Table 1.** Cohort characteristics, data presented as median (IQR), as data was non-normally distributed. Symptom duration is reported from symptom onset to day of study enrolment (February 2022 for long COVID and October 2023 for ME/CFS).

|  | Bed Rest |  | Control | Long COVID | ME/CFS |
| --- | --- | --- | --- | --- | --- |
|  | Pre | Post |  |  |  |
| **Participants** | 24 | 24 | 30 | 25 | 26 |
| **(# Females)** | (8) | (8) | (15) | (13) | (13) |
| **Age** | 30 | 30 | 42 | 43 | 42 |
| **(years)** | (27-38) | (27-37) | (33-54) § | (31-50) § | (32-50) § |
| **Height** | 175 | 175 | 179 | 177 | 176 |
| **(cm)** | (172-181) | (170-181) | (174-186) | (171-184) | (171-186) |
| **Weight** | 74 | 74 | 76 | 80 | 84 |
| **(kg)** | (70-79) | (69-79) | (64-88) | (69-92) | (75- 89) |
| **Symptom Duration** | n/a | n/a | n/a | 545 | 3793 |
| **(days)** |  |  |  | (455-686) | (2510-5496) |
|  |  |  |  | ††† |  |
| **Daily Steps** | ND | 0 | 7153 | 4718 | 3704 |
| **(steps\*day<sup>-1</sup>)** |  |  | (5063-8405) | (2794-5832) | (2286-5052) |
|  |  |  | ††† | *** | *** |

\*: vs. CON; †: vs. ME/CFS; §: vs. BED REST
\*: p<0.05; \*\*: p<0.01; \*\*\*: p<0.001

Healthy bed rest participants were on average younger than healthy controls matched to patients (*P*=0.02). In a sub-group analysis, we age- and sex-matched pre-bed rest participants to patient-matched healthy controls. 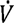*0*_2*max*_ (Supplemental Figure 1A) and the gas exchange threshold (GET; Supplemental Figure 1B-C) did not differ between cohorts. Fiber cross sectional area (FCSA; Supplemental Figure 1D) was also similar between groups, however, the pre-bed rest cohort exhibited lower capillary-to-fiber ratios (Supplemental Figure 1E), lower succinate dehydrogenase activity (Supplemental Figure 1F), and a tendency to have a lower oxidative phosphorylation capacity (*P*=0.068, Supplemental Figure 1G). As some skeletal muscle features differed between those control groups, we analyzed them separately: healthy controls were matched to patients with ME/CFS and long COVID (Table 1), whereas the long-term bed rest cohort were analyzed longitudinally.

### Acute Exercise Responses

Bed rest is known to reduce maximal aerobic exercise capacity and we expected both patient groups to exhibit lower maximal aerobic exercise capacities compared to healthy controls. Indeed, maximal oxygen uptake (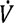*0*_2*max*_; Figure 1B) and peak power output (Supplemental Figure 2A) were reduced after 60-day bed rest, and were similarly lower in patients with long COVID and ME/CFS compared to healthy controls (Figure 1B + Supplemental Figure 2A).

The GET, a submaximal non-invasive marker for the lactate threshold, was reduced following bed rest (Figure 1C), and was similarly lower in both patient groups compared to healthy controls (Figure 1C). Following bed rest, GET occurred at a higher percentage of 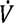*0*_2*max*_ (Supplemental Figure 2B), whereas in patients with ME/CFS, GET occurred at a lower percentage of 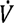*0*_2*max*_ compared to both patients with long COVID and healthy controls (Supplemental Figure 2B), suggesting an earlier relative onset of lactate accumulation in patients.

Bed rest also altered ventilatory and cardiovascular responses to exercise. Maximal minute ventilation was reduced (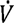*E_max_*; Figure 1D), whereas ventilatory equivalents for O_2_ and CO_2_ (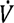*E_max_*/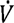*C0*_2*max*_ and 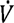*E_max_* /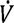*0*_2*max*_), and the 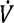E/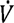 *C0*_2_ slope (Figure 1E + Supplemental Figure 2C-D) increased. Cardiovascular alterations included a reduced maximal O_2_-pulse (i.e. 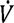*0*_2_/heart rate, equal to the product of stroke volume and arteriovenous O_2_ difference; Supplemental Figure 2E); increased maximal heart rate (Supplemental Figure 2F), and increased adjusted heart rate reserve (AHRR; Supplemental Figure 2G).

While 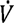*E_max_* (Figure 1D) tended to be lower in patients compared to healthy controls (*P*=0.056), there were no significant differences in 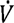*E_max_*/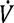*C0*_2*max*_, 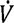*E_max_*/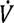*0*_2*max*_, the 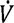E/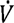 *C0*_2_ slope, maximal heart rate, or AHRR (Figure 1E, Supplemental Figure 2C+D+F+G). However, both patients with long COVID and ME/CFS exhibited decreased O_2_-pulse compared to healthy controls (Supplemental Figure 2E), similar to the effects of bed rest. Patients with ME/CFS exhibited a more pronounced increase in heart rate relative to oxygen uptake (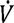*0*_2_-HR slope; Supplemental Figure 2H) compared to healthy controls, a pattern not seen following bed rest.

### Skeletal Muscle Fiber Type Alterations

As muscle unloading can induce atrophy within days^22^, we assessed muscle fiber cross sectional area (FCSA), fiber-type specific FCSA, and fiber type composition using immunohistochemistry (Figure 2A). As expected, bed rest induced muscle atrophy (Figure 2B), affecting all fiber types similarly (Supplemental Figure 2A). In contrast, overall muscle FCSA of patients with long COVID and ME/CFS did not differ from healthy controls (Figure 2B). However, when assessing fiber-type specific FCSA, patients with ME/CFS exhibited significantly smaller type I fibers compared to healthy controls (Supplemental Figure 3B), while all other fiber type sizes remained similar to healthy controls. This suggests selective atrophy of type I fibers in patients with ME/CFS but not after bed rest.

**Figure 2:**
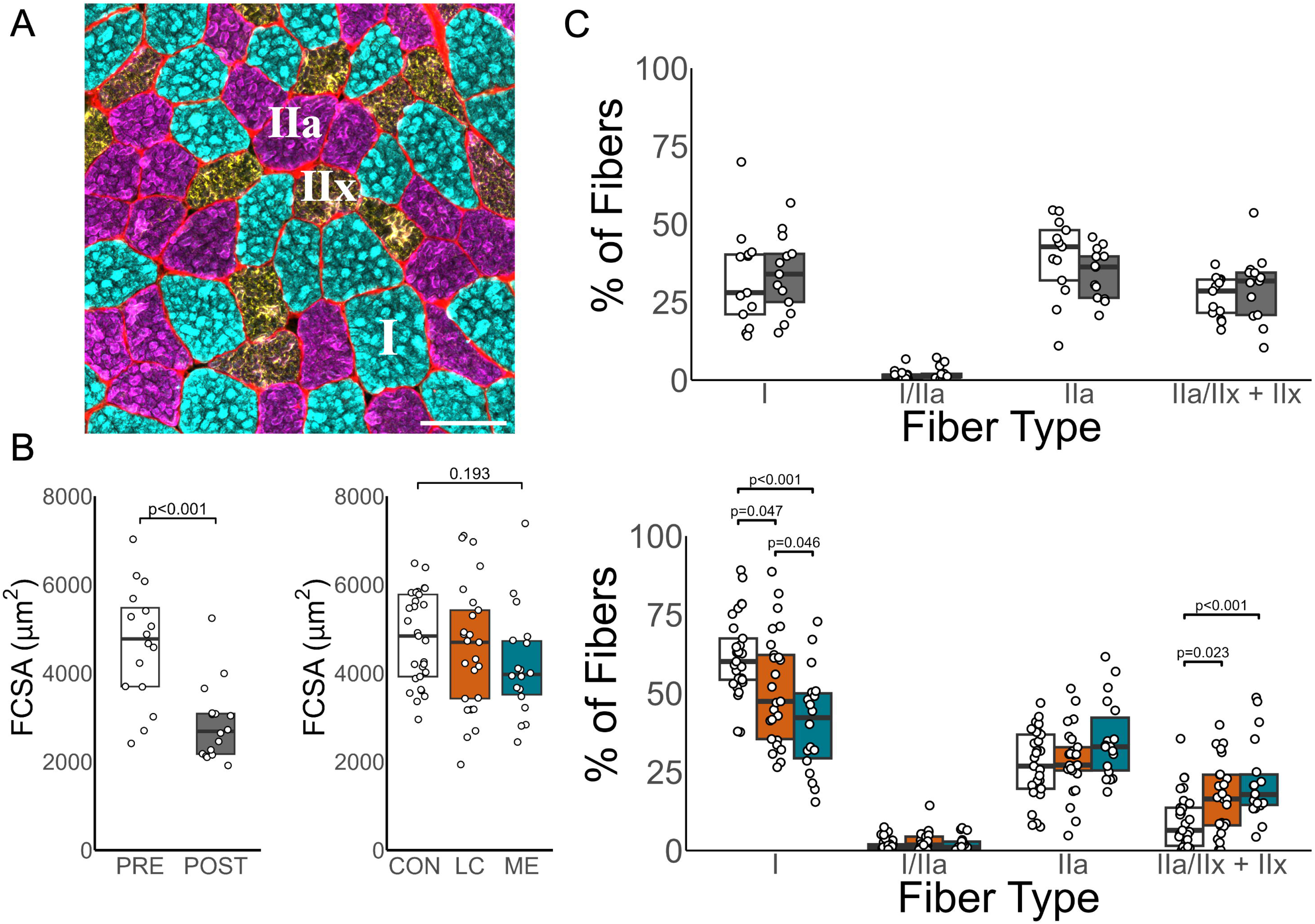
**A:** A typical example of muscle fiber type staining with colours adjusted to be colour-blind visible. Scale bar represents 100µm. Severe atrophy was observed following bed rest (**B**), whereas fiber cross-sectional area (FCSA) was not different between patients with long COVID and myalgic encephalomyelitits/chronic fatigue syndrome (ME/CFS) and healthy controls (**B**). Following bed rest, fiber type composition (**C**) was not significantly altered. Long COVID and ME/CFS patients exhibited lower proportions of type I fibers compared to healthy controls, while both patients with Long COVID and ME/CFS had significantly higher proportions of type IIa/IIx+IIx fibers compared to healthy controls. All data was normally distributed following Box-Cox transformations, except proportions of IIa/IIx+IIx fibers. Paired t-tests were used to assess parametric data from the bed rest cohort. Non-parametric data from the bed rest cohort was assessed using Mann–Whitney *U* test. ANOVA, with Tukey HSD post-hoc testing or Kruskal–Wallis *H* test, with pairwise Wilcoxon tests with Benjamini–Hochberg correction post-hoc for patient cohorts.

Following bed rest, fiber type composition did not change (Figure 2B). In contrast, patients with long COVID and ME/CFS had significantly lower proportion of type I fibers and a greater proportion of type IIa/IIx + IIx fibers compared to healthy controls (Figure 2C), while patients with ME/CFS had an even lower proportion of type I fibers than patients with long COVID.

### Mitochondrial Respiration and Activity

Given the association between mitochondrial function and whole-body aerobic exercise capacity^29^, we assessed mitochondrial respiration of permeabilized fibers and succinate dehydrogenase (SDH) activity in sections, to provide further insight into mitochondrial alterations upon bed rest and in patients. Bed rest resulted in lower SDH activity and oxidative phosphorylation capacity (Figure 3A+B). Compared to healthy controls, patients with ME/CFS exhibited lower SDH activity (Figure 3A), whereas patients with long COVID were not different to healthy controls. Both patients with long COVID and ME/CFS presented with lower oxidative phosphorylation capacities compared to healthy controls (Figure 3B). Both SDH activity and oxidative phosphorylation capacity were correlated with 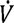*0*_2*max*_ in healthy controls (Figure 3C+D), and was present following bed rest. However, this association was absent in patients with long COVID and ME/CFS. The lack of association of 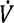*0*_2*max*_ with mitochondrial variables suggests that their reduced exercise capacity is not solely explained by mitochondrial respiration or enzymatic activity.

**Figure 3:**
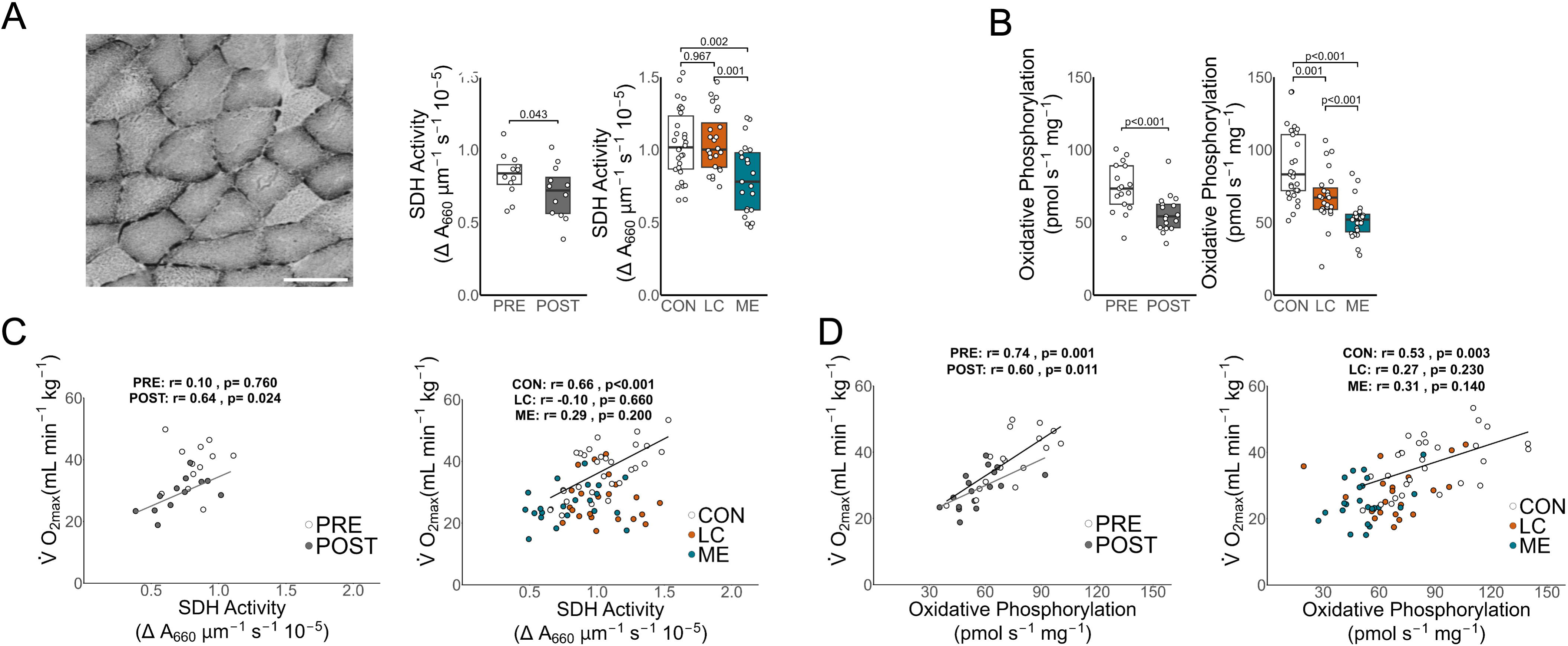
Succinate dehydrogenase (SDH) activity (**A**) was significantly reduced following bed rest. SDH activity in patients with Long COVID was not different to healthy controls but lower in ME/CFS. Oxidative phosphorylation capacity (**B**) was reduced following bed rest and was lower in both patients with long COVID and ME/CFS compared to healthy controls. SDH was associated with 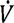*O*_2*max*_ (**C**) after bed rest and in healthy controls. The relationship between oxidative phosphorylation capacity and 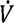*O*_2*max*_ (**D**) was significant in pre- and post-bed rest and healthy controls, but not in patients with ME/CFS or long COVID. Bars represent 100µm. Oxidative phosphorylation data in the patient cohort remained non-normally distributed following Box-Cox transformation. Paired t-tests were used to assess parametric data from the bed rest cohort. Non-parametric data from the bed rest cohort was assessed using Mann–Whitney *U* test. ANOVA, with Tukey HSD post-hoc testing or Kruskal–Wallis *H* test, with pairwise Wilcoxon tests with Benjamini– Hochberg correction post-hoc for patient cohorts. Linear relationships were assessed using Pearson’s correlation, solid lines represent significant correlations (*P*<0.05).

As mitochondrial respiration is a reflection of both mitochondrial content and function, we subsequently normalized mitochondrial respiration to gain insight to alterations of intrinsic mitochondrial function. After normalizing mitochondrial respiration to SDH activity, there was no effect of bed rest on oxidative phosphorylation capacity (Supplemental Figure 4A). In contrast, patients with long COVID still displayed lower oxidative phosphorylation capacity normalized to SDH activity compared to healthy controls, and patients with ME/CFS tended to exhibit lower oxidative phosphorylation capacity (*P*=0.063, Supplemental Figure 4A). Normalizing maximal uncoupled respiration to leak respiration (E/L coupling efficiency) is reflective of intrinsic mitochondrial biochemical coupling, wherein a less coupled system (lower values) indicates higher proton leak. Bed rest did not alter the E/L coupling efficiency, however patients tended to display lower E/L coupling efficiency compared to healthy controls (Supplemental Figure 4B). Bed rest increased the NADH-linked flux control ratio (PN/PNS) and deceased the succinate-linked flux control ratio (PS/PNS, Supplemental Figure 4C-D), whereas there was no difference amongst patients and healthy controls.

### Skeletal Muscle Capillarization

As bed rest induced muscle fiber atrophy and patients exhibited a lower proportion of type I fibers, we hypothesized that capillary supply may be differentially affected. Following bed rest, the average capillary-to-fiber ratio was unchanged (Figure 4A), however capillary cross sectional area was significantly reduced (Supplemental Figure 5A). Due to greater muscle atrophy relative to capillary loss^22^, capillary density was increased following long-term bed rest (Figure 4C). However, these adaptations were not observed in patients with long COVID or ME/CFS. Patients with ME/CFS displayed lower capillary-to-fiber ratios and capillary densities (Figure 4C) compared to healthy controls and patients with long COVID, while capillary measures in patients with long COVID were not different from healthy controls. The relationship between fiber size and the capillary-to-fibre ratio is reflective of adaptive remodelling to support diffusive oxygen supply in larger fibers. In all groups, capillary-to-fiber ratios were proportional to FCSA, except following long-term bed rest, where the relationship trended towards significance (*P*=0.062; Figure 4D). However, both patient groups exhibited significantly lower intercepts than healthy controls (both *P*<0.05), indicating less capillary supply for a given fiber cross sectional area. This may impair diffusive oxygen supply, however, whether compensatory adaptations in intracellular myoglobin content occur remained unclear. To this end, we evaluated the intracellular myoglobin content in a subset of participants. Following bed rest, myoglobin content increased unexpectedly (Supplemental Figure 6A), whereas patients and healthy controls did not differ.

**Figure 4:**
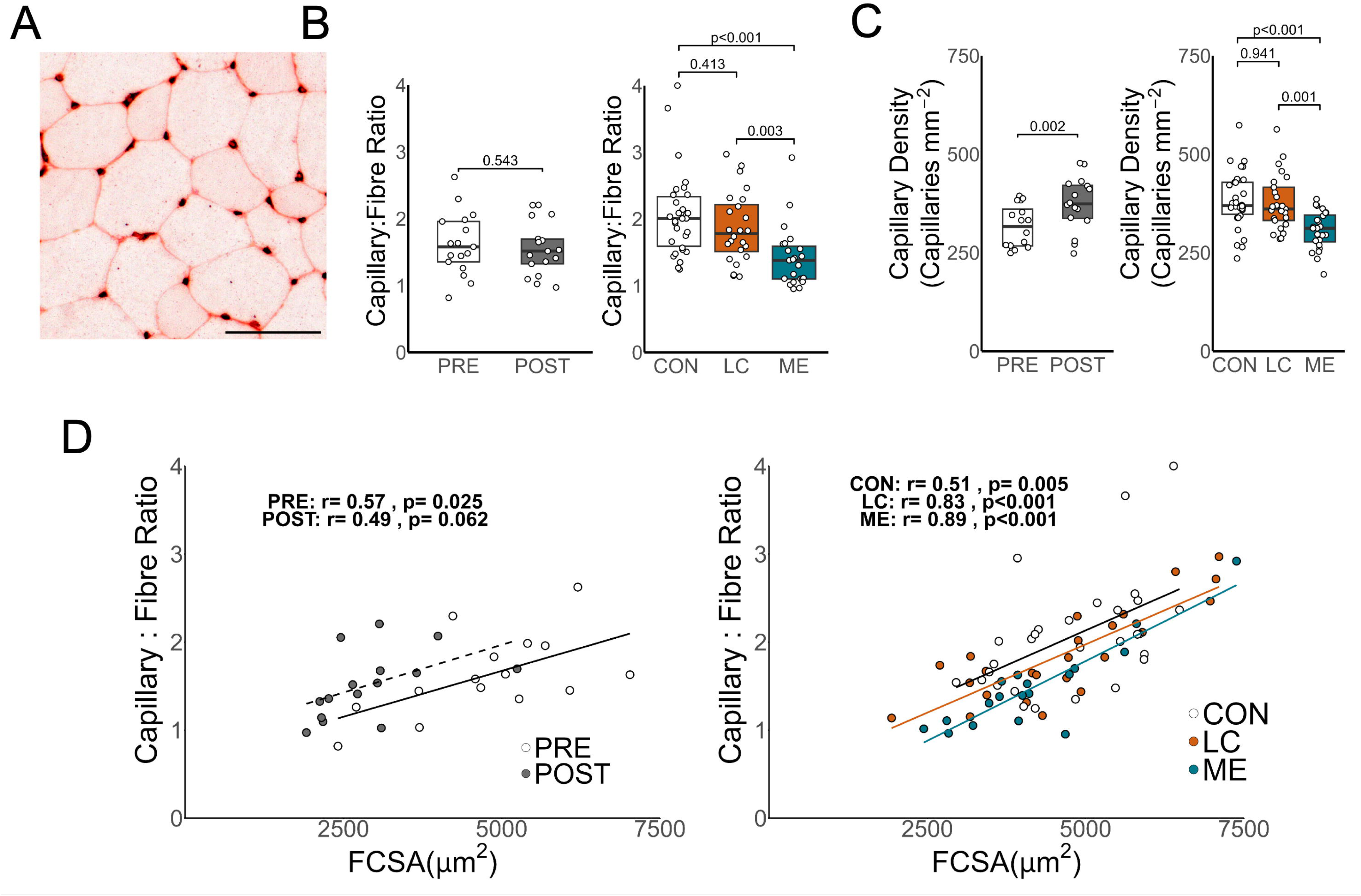
A typical example of a lectin stain for capillaries is shown in **A**. Scale bar represents 100µm. Following bed rest, capillary-to-fiber ratio was not significantly different (**B**), while patients with ME/CFS had a significantly lower capillary-to-fiber ratio compared to patients with long COVID and healthy controls. Capillary density (**C**) was significantly increased following bed rest, while both patients with long COVID and patients ME/CFS had significantly lower capillary densities compared to healthy controls (**D**). The relationship between fiber cross-sectional area (FCSA) and capillary-to-fiber ratio significantly correlated in all conditions (albeit *P*=0.062 following bed rest). The relationship between FCSA and capillary-to-fiber ratio was not significantly different following bed rest (z=0.495, *P*=0.621), however compared to healthy controls, both patients with long COVID (z= - 2.176, *P*=0.030) and patients with ME (z= −2.602, *P*=0.009) displayed significantly different relationships (**G**). Capillary density and capillary-to-fiber ratio data were normally distributed following Box-Cox transformation. Paired t-tests were used to assess parametric data from the bed rest cohort. Non-parametric data from the bed rest cohort was assessed using Mann–Whitney *U* test. ANOVA, with Tukey HSD post-hoc testing or Kruskal–Wallis *H* test, with pairwise Wilcoxon tests with Benjamini–Hochberg correction post-hoc for patient cohorts. Linear relationships were assessed using Pearson’s correlation on non-transformed data. Solid lines represent significant correlations, dashed lines represent correlations with *P*<0.10.

### Comparing skeletal muscle alterations in long COVID and ME/CFS with long-term bed rest

Since both patient groups had lower daily physical activity levels than healthy controls, we reasoned that healthy participants following bed rest could serve as an alternative control group. We therefore analyzed a subgroup of 13 patients with ME/CFS, 13 patients with long COVID and 13 age- and sex-matched healthy individuals following long-term bed rest (participant characteristics outlined in Table S1).

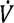*0*_2*max*_ did not differ between groups (Supplemental Figure 7A), however patients with ME/CFS had significantly lower GET than long COVID (Supplemental Figure 7B), with healthy post-bed rest individuals displaying intermediate values. GET occurred at a lower relative intensity in patients with ME/CFS than in both patients with long COVID and participants following bed rest (Supplemental Figure 7C).

Participants after bed rest had significantly smaller FCSA compared to both patient groups (Supplemental Figure 7D). Patients with long COVID displayed higher capillary-to-fiber ratios and SDH activity than participants after bed rest and patients with ME/CFS (Supplemental Figure 7E+F). Patients with long COVID displayed higher oxidative phosphorylation capacity compared to patients with ME/CFS, and also tended to be higher than participants after bed rest (Supplemental Figure 7G, *P*=0.061).

When assessing the relationships between capillary-to-fiber ratio and FCSA, only patients with long COVID displayed a significant relationship, while patients with ME/CFS showed a similar trend (Supplemental Figure 7H). Conversely, only participants after bed rest displayed a significant correlation between 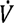*0*_2*max*_ and SDH activity (Supplemental Figure 7I). These results suggest that capillary supply and mitochondrial function are disrupted differently in long COVID and ME/CFS compared to bed rest-induced deconditioning.

## Discussion

The major finding of the present study is that acute exercise responses and skeletal muscle alterations in long COVID and ME/CFS differ from those caused by bed rest-induced deconditioning per se. Despite similar whole body exercise capacity in both patient groups and participants after bed rest, the pulmonary and cardiovascular responses to acute exercise were distinct. Bed rest induced severe muscle atrophy, which was not observed in patients with long COVID or ME/CFS. Instead, both patient groups displayed a lower proportion of type I fibers, whereas only patients with ME/CFS exhibited lower capillary-to-fiber ratios and type I specific atrophy. Further, the lower oxidative phosphorylation capacity in both patient groups did not correlate with 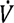*0*_2*max*_, in contrast to bed rest participants, suggesting that physiological impairments distinct from inactivity drive reduced exercise capacity in patients. These findings challenge the notion that deconditioning is the primary driver of reduced exercise capacity in long COVID and ME/CFS. Rather, our observations suggest intrinsic skeletal muscle abnormalities contribute to limited exercise capacity, including reduced capillary supply relative to muscle fiber size and mitochondrial impairments.

### Exercise Responses

Maximal aerobic capacity was similar between patients with long COVID and ME/CFS, and those undergoing a strict 60-day bed rest. However, the physiological determinants underpinning this reduction differed. Ventilatory responses to acute exercise differed following bed rest compared to the patient groups. Bed rest resulted in reduced 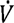*E_max_*, consistent with the decrease in 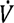*0*_2*max*_, whereas the 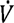*E_max_*/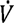*C0*_2*max*_ and the 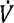*E*-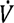*C0*_2_ slope were increased. Patients only exhibited higher 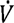*E*/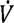*C0*_2_ during submaximal exercise, and a tendency of lower 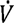*E_max_*. These findings suggest that bed rest and both disease conditions induce ventilatory inefficiencies characterized by marked hyperventilation during submaximal exercise. The causes of these findings are currently unclear, however reduced blood or muscle CO_2_ buffering capacity^30^, increased chemoreceptor sensitivity^31,32^, pulmonary vascular dysfunction resulting in increased dead space ventilation^33^, or respiratory muscle fatigue^34,35^ may contribute.

Acute cardiovascular responses to exercise also differed between cohorts. Previous studies report that deconditioning typically leads to an increase of the 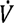*0*_2_-heart rate slope^23^, reflecting a greater heart rate increase per unit change in 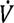*0*_2_, which is often associated with reduced oxygen extraction^23,36,37^. However, in our study, bed rest participants did not present with this pattern, which may be attributed to the disproportionate increase in baseline heart rate (+30%) relative to their maximal heart rate (+5%) and reduction in 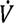*0*_2*max*_ (−24%) following bed rest. In contrast, our patient groups exhibited an increased 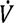*0*_2_-heart rate slope, suggesting a greater reliance on changes in heart rate to satisfy a given oxygen demand. While this may be indicative of either decreased stroke volume or reduced peripheral oxygen extraction^23^, it is consistent with results from invasive CPET studies, which have shown marked reduction in peripheral oxygen extraction in long COVID and ME/CFS patients^38,39^.

Future studies in long COVID and ME/CFS should focus on measuring markers of cardiovascular function during exercise, such as stroke volume, peripheral oxygen extraction, and baroreflex sensitivity to further elucidate the underlying mechanism.

### Skeletal Muscle Oxidative Changes

The reduction in oxidative phosphorylation capacity following bed rest was shown to be primarily attributed to a reduction in SDH activity, often used as a proxy for mitochondrial density^18,19,40–42^. In contrast, the impaired oxidative phosphorylation capacity in both patient groups was still present following normalization to SDH activity, suggesting that intrinsic mitochondrial respiration may be primarily affected. Furthermore, while E/L coupling efficiency tended to be reduced in both patient groups, this was not observed following bed rest. These results suggest that skeletal muscle oxidative impairments in long COVID and ME/CFS cannot solely be attributed to deconditioning. Indeed, previous work suggests abnormal ultrastructure, particularly reduced cristae density, in patients with long COVID and ME/CFS^11^, while bed rest is typically associated with more fragmented mitochondria^18^. What the cause of the intrinsic mitochondrial alterations in long COVID and ME/CFS is, is currently unclear. Future work is required to quantify skeletal muscle mitochondrial ultrastructure in long COVID and ME/CFS.

Notably, oxidative phosphorylation capacity and SDH activity correlated with whole-body maximal oxygen uptake in healthy individuals (before and after bed rest), whereas these were absent in long COVID and ME/CFS. This suggests that factors beyond skeletal muscle mitochondrial respiration constrain exercise capacity in long COVID and ME/CFS, including impairments in endothelial function^43–47^, a reduced venous return and cardiac preload^38^, or a reduction in muscle diffusive capacity^48^ contributing to impaired peripheral oxygen extraction^39^. Conversely, mitochondrial respiration has been shown to constrain 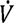*0*_2*max*_ in healthy sedentary, untrained individuals^49,50^, which is consistent with the strong correlations observed between mitochondrial markers and 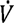*0*_2*max*_ across all healthy cohorts. Thus, it is likely the constraints on 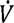*0*_2*max*_ imposed by bed rest, long COVID and ME/CFS differ, implying that the reduced aerobic exercise capacity in long COVID and ME/CFS cannot solely be ascribed to deconditioning.

### Capillary network and myoglobin content

Capillary-to-fiber ratios and capillary density were lower in patients with ME/CFS, but not long COVID, compared to healthy controls, reducing the ability to adequately supply oxygen and nutrients, and remove metabolic byproducts during exercise. These findings are contrary to previous reports of lower capillary densities in long COVID^9,51^. In contrast, while capillary-to-fiber ratio was unaltered following bed rest, capillary density was increased due to significant muscle atrophy^22,42^, consistent with the principle that alterations in muscle size typically occur before capillary alterations^22,52,53^. Intriguingly, the average area of individual capillaries was reduced following bed rest (Supplemental Figure 5A). This may be due to reduced capillary tortuosity^54,55^ resulting in more obliquely cut capillaries, or due to the combination of lower venous pressure, the lack of hydrostatic pressure and shear stress experienced during head-down tilt resulting in a structural remodeling of the capillary beds^56^. That skeletal muscle microvascular-endothelial function was impaired following a 10-day bed rest^57^ is consistent with this notion. Although we report unaltered structural indices of capillarization in long COVID, it is unknown whether skeletal muscle capillary perfusion (and oxygen diffusion) was altered during exercise in either long COVID or ME/CFS, as direct measurements of skeletal muscle capillary blood flow are challenging to assess in humans *in vivo*.

Capillary-to-fiber ratio is highly related to muscle FCSA, wherein larger fibers typically have more capillary contacts to accommodate oxygen and nutrient supply, and metabolite removal^52,53,58^. While these relations existed in all groups, except following bed rest, this relationship was downshifted in both long COVID and ME/CFS patients compared to healthy controls. This suggests that less capillaries are available to supply oxygen and nutrients for a given FCSA in patients, which is distinct from the capillary alterations following strict bed rest^22^. The reduced capillary-to-fiber for a given muscle size may result in local tissue hypoxia at high oxygen utilization rates (i.e. maximal exercise) in patients with long COVID and ME/CFS. Importantly, the lower maximal mitochondrial respiration reduces local oxygen utilization rates in skeletal muscle, Tissue hypoxia markers are technically difficult to assess, and is currently unknown if skeletal muscle hypoxia-inducible factor-1α (HIF1α) signaling is altered in long COVID or ME/CFS.

To study downstream markers of local tissue hypoxia, we assessed myoglobin content in the skeletal muscle cross sections (Supplemental Figure 5B). We suspected that if local oxygen supply was reduced, there would be a compensatory increase in myoglobin content, and likewise, oxygen extraction was impaired, myoglobin content would be reduced. However, myoglobin content in neither patients with long COVID nor ME/CFS differed from healthy individuals. In contrast, bed rest increased myoglobin content (Supplemental Figure 5B)^59^, suggesting either reduced oxygen perfusion^57^ or decreased oxygen extraction^37,40,60^ following bed rest. In conclusion, we do not find evidence of local hypoxia inside skeletal muscle in patients with long COVID and ME/CFS, likely because the ratio between local oxygen supply and utilization rates are maintained.

### Muscle Fiber Type and Size

We confirmed that bed rest causes significant atrophy across all fiber types^25,61,62^. Conversely, overall FCSA in patients with long COVID ME/CFS was not different from healthy controls. However, patients with ME/CFS exhibited selective type I fiber atrophy (Supplemental Figure 3A), suggesting fiber-type specific vulnerability to atrophy. The underlying mechanism remains unclear, as we lack the longitudinal data to determine whether these changes result from the disease progression or pre-disease differences.

Previous studies show conflicting results regarding changes in fiber type composition following bed rest^22,63–66^. In our cohort, fiber type composition remained unchanged following bed rest. In contrast, both patients with long COVID and ME/CFS exhibited higher proportions of type IIa/IIx fibers and lower proportions of type I fibers compared to healthy controls. Since we did not obtain longitudinal muscle biopsies from patients, we cannot determine whether this is due to a fiber type transition, or whether individuals with higher proportions of type IIa/IIx fibers are predisposed to developing long COVID or ME/CFS. However, given the type I fiber-specific atrophy pattern observed in patients with ME/CFS, the lower proportion of type I fibers and higher proportions of type IIa/IIx fibers in patients, our findings suggest that a shift towards type IIa/IIx fibers may occur in ME/CFS. The atrophy and the fiber type shifts were more pronounced in ME/CFS compared to long COVID patients, possibly because ME/CFS patients were diagnosed before the COVID pandemic, and therefore were ill for a significantly longer time. Whether the disease duration or intrinsic differences between long COVID and ME/CFS underlie these differences is therefore unknown.

### Limitations

The current study indicates that skeletal muscle adaptations and lower aerobic exercise capacities observed in patients with long COVID and ME/CFS are different from those occurring after bed rest, however there are several limitations. We did not induce physical inactivity in patients with ME/CFS or long COVID. Patients are typically more sedentary because PEM worsens both their symptoms and skeletal muscle abnormalities^6^. We also cannot completely exclude the contribution of physical inactivity to the skeletal muscle alterations observed in patients with long COVID and ME/CFS, however our results do indicate that physical inactivity alone is insufficient to explain the skeletal muscle changes in patients. There was likely self-selection bias towards patients with milder symptoms, as they volunteered to travel to the laboratory for multiple occasions and undergo a maximal exercise test. Our results are therefore not directly applicable to home-bound patients ^67^.

Lastly, the current research design precludes us to draw conclusions about the differences in pathophysiology of long COVID and ME/CFS. ME/CFS patients were ill for a significantly longer period, and therefore we cannot distinguish between disease duration and disease pathophysiology.

### Conclusions

This study demonstrates that the skeletal muscle determinants of exercise capacity in long COVID and ME/CFS differ from those induced by prolonged bed rest. While 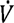*0*_2*max*_ and markers for aerobic exercise capacity were similarly low in patients with long COVID and ME/CFS compared to long-term bed rest, respiratory and cardiovascular responses to acute exercise were distinct. Patients with long COVID and ME/CFS displayed higher proportions of type IIa/IIx fibers, and signs of intrinsic mitochondrial dysfunction, observations that were not seen following bed rest. These findings indicate that the lower exercise capacity in patients with long COVID and ME/CFS is not solely due to physical inactivity; therefore rehabilitation strategies for patients should consider that patients with ME/CFS and long COVID are not simply deconditioned, and should be treated as unique cases with alternative rehabilitation strategies.

## Methods

### Study Approval

Two cohorts were compared: one consisting of 24 bed rest participants undergoing a strict 60-day head-down tilt bed rest (occurring before 2020), and one consisting of a cross-sectional design cohort of 25 patients with long COVID, 26 patients with ME/CFS and 30 age- and sex-matched healthy controls that had successfully recovered from an acute SARS CoV-2 infection. Participants undergoing bed rest were part of the AGBRESA study (registered at DRKS00015677)^22^. The protocol was approved by the ethics commissions of the Medical Association North Rhine (number 2018143) and NASA (Johnson Space Center, Houston, United States). Patients with long COVID and patients with ME/CFS were part of a case-control study in the Amsterdam University Medical Centers (AUMC) and the Faculty of Behavioral and Movement Sciences (Vrije Universiteit Amsterdam). The study protocol was approved by the medical ethics committee of the Amsterdam UMC (NL78394.018.21) and registered at www.clinicaltrials.gov (NCT05225688). All participants signed a written informed consent before participation. The study was conducted in accordance with the Declaration of Helsinki. Some data have previously been published^6^.

### Study populations

#### Bed rest

The study details have been reported previously^22^. Briefly, 24 healthy participants underwent a 60-day strict 6 degrees head-down tilt bed rest (Table 1). Participants followed standardized diets, consuming 1.6 times their resting metabolic rate before bed rest and 1.3 times their resting metabolic rate during bed rest. Fluid intake was controlled, and ingestion of caffeine and alcohol were prohibited.

### Long COVID and ME/CFS cohort

Patients with long COVID were diagnosed by two experienced clinicians for long COVID symptomology and exclusion of potential differential diagnoses. All long COVID patients were diagnosed with post-exertional malaise (PEM) by the DSQ-PEM^68^, had a minimum period of long COVID-related symptoms of six months, and were between 18 and 65 years old. No symptoms were present before the confirmed diagnosis of SARS-CoV-2. None of the included participants were admitted to the hospital during acute SARS-CoV-2 infection and were healthy prior to NAAT or serology-proven SARS-CoV-2 infection. Reported symptom durations (Table 1) were from symptom onset to study enrolment (February 2022 for long COVID and October 2023 for ME/CFS). Exclusion criteria were a medical history of cardiovascular/pulmonary disease, diabetes mellitus, concurrent treatment with metabolism or coagulant-altering drugs during the study period (statins, corticosteroids, SGLT2 inhibitors, GLP1 receptor agonists, platelet aggregation blockers and any anticoagulants), adiposity and body mass index >35 (related to muscle biopsy obtainment), pregnancy, active infection, severe renal dysfunction or any other prior chronic illness or >6 alcohol units per day or >14 alcohol units per week. Smoking was not an exclusion criterion. One long COVID patient was excluded due to a recent SARS-CoV-2 re-infection (<7 days).

Patients with ME/CFS all fulfilled the Canadian Consensus Criteria (CCC), exhibited PEM according to the DSQ-PEM and consulted with a ME/CFS specialist or post-COVID physician, and were between 18 and 65 years old. ME/CFS diagnosis was prior to 2020, and all patients had a confirmed previous diagnosis of SARS-CoV-2. Exclusion criteria were identical to those of the patients with long COVID.

None of the healthy controls had residual symptoms after the SARS-CoV-2 infection and none were hospitalized within 6 months of study participation. Healthy controls withdrew because of the invasive nature of the protocol (*n*□=□2), symptoms related to a burn-out (*n*□=□1), and a novel diagnosis of uncontrolled hypertension (*n*□=□1).

### Cardiopulmonary Exercise Testing

All participants underwent cardiopulmonary exercise testing (CPET) on an electronically braked cycle ergometer to assess aerobic exercise capacity. Generally, each test was preceded rest on the ergometer, followed by a baseline cycling phase before beginning the incremental exercise test.

Bed rest participants performed a step incremental test (Lode Excalibuer, Lode, Groningen, The Netherlands), whereas patients with long COVID and ME/CFS performed a ramp incremental exercise test (Lode Excalibur Sport, Lode, Groningen, The Netherlands; Monark L7TT, Monark Sports & Medical, Vansbro, Sweden). Step and ramp protocols have been reported to yield comparable 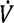*0*_2*max*_ values^69^.

#### Step Test Protocol (bed rest cohort)

Participants first completed 5-min of quiet rest on the cycle ergometer, followed by 3-min of baseline cycling at 50W. Power output then increased by 25 W min^-1^ until task failure. Participants were instructed to maintain 75 RPM, and task failure was defined as the point at which cadence dropped below 70 RPM despite verbal encouragement.

#### Ramp incremental test (Long COVID & ME/CFS cohort)

Participants completed 2-min quiet rest on the ergometer and 4-min baseline cycling at low intensity between 0 and 20□W. This was followed by a ramped, linear increase in power output (10-60W min^-1^) until task failure. The individual baseline work rates and ramp slopes were selected based on each participant’s anthropometric characteristics and physical activity levels and designed to elicit task failure within 8–12□min. Participants were instructed to maintain their cadence between 70 and 90 revolutions/min, and task failure was defined as the point at which cadence dropped below 60 revolutions/min despite verbal encouragement. Capillary lactate concentrations were determined at rest prior to the onset of the test, during baseline cycling, and immediately following task failure (Lactate Pro 2 LT-1730, ARKRAY Ltd., United Kingdom). Pulmonary gas exchange and ventilation were measured on a breath-by-breath basis (Bed rest: Innocor, Innovision, Odense, Denmark; Patients with long COVID and patients with ME/CFS: Cosmed Quark CPET; Cosmed, Rome, Italy). In all studies, 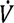*0*_2*max*_ was defined as the highest 30 second rolling average 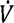*0*_2_ achieved during the test. Achievement of 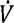*0*_2*max*_ was determined by the presence of a plateau of 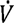*0*_2_ prior to cessation of exercise^70^.

The gas exchange threshold (GET) was assessed using 10-second binned averages as the point of non-linear rise in 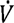*C0*_2_ relative to 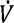*0*_2_ using the V-slope method^71^, and verified with visual inspection. The GET was then confirmed visually by assessing 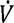*E* vs 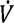*0*_2_, 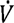*E*/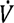*C0*_2_ vs 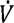*0*_2_, and 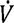*E*/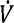*0*_2_ vs 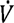*0*_2_. Respiratory compensation point (RCP) was assessed as the point of non-linear rise in 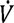*E* relative to 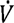*0*_2_ using the V-slope method, and again verified visually. Presence of the RCP was confirmed by visual inspection of the 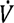*E*/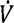*C0*_2_ vs 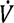*0*_2_ and PetCO_2_ vs 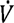*0*_2_.

Maximal acute exercise values were taken as the highest 30-second rolling averages of the respective parameters. The 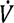*E*/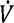*C0*_2_ slope was calculated as the linear rise until RCP. 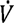*0*_2_-heart rate slopes were calculated using the entirety of the exercise test and taken as the slope of heart rate compared to 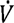*0*_2_. Baseline heart rate was calculated using an average over the 5-minute quiet rest, with the first 30-seconds and last 30-seconds being disregarded. Adjusted heart rate reserve was calculated as the (HR_max_-HR_baseline_)/(220-age-HR_baseline_)^72^.

Due to software issues, three patients with ME/CFS undertook ramp rates that were too high, resulting in a duration of exercise that was not sufficient to establish acute exercise responses. Evaluation of maximal exercise was still possible in these participants, thus submaximal values were excluded.

### Skeletal muscle measurements

#### Biopsy Procedure

Muscle biopsies were obtained from the vastus lateralis at one-third of the distal length (approximately 150mg weight wet) using a rongeur (4mm diameter for the bed rest cohort) or a suction-supported 5mm Bergström needle (Pelomi, Albertslund Denmark; for the Long COVID & ME/CFS patient cohort) under sterile conditions after skin disinfection and local anaesthesia with 2% lidocaine^6,18^. Muscle biopsies were taken before and after 55 day of head-down tilt bed rest. One ∼30 mg piece was aligned according to the fiber arrangement under a light microscope and frozen in liquid nitrogen for histochemistry, and another ∼15 mg piece was utilized for high-resolution respirometry experiments.

All biopsies used for histochemistry were frozen in liquid nitrogen and stored in liquid nitrogen at −196°C. Frozen muscle samples were mounted using freeze-gel (Q Path, VWR, Netherlands) and cut into 10 µm thick sections in transverse orientation in a cryostat (Microm HM550, Adamas Instrumenten, Netherlands) at −20°C, before being mounted on polylysine-coated slides, and stored at −80°C until staining. Due to issues with tissue freezing, histological analysis was completed on samples from 17 participants pre- and post-bed rest, 24 patients with long COVID, 23 patients with ME/CFS, and 30 healthy controls.

### Skeletal muscle respirometry

Mitochondrial respiration was assessed in permeabilized fibers as described previously^6^. Small bundles of freshly isolated fibers were permeabilized with 50□µg□mL^−1^ saponin for 20□min at 4□°C in a solution consisting of (in mM) CaEGTA (2.8), EGTA (7.2), ATP (5.8), MgCl_2_ (6.6), taurine (20), phosphocreatine (15), imidazole (20), DTT (0.5) and MES (50) (pH 7.1). Tissue was washed in respiration solution containing EGTA (0.5), MgCl_2_ (3), K-lactobionate (60), taurine (20), KH_2_PO_4_ (10), HEPES (20), sucrose (110) and 1□g□L^−1^ fatty acid-free BSA (pH 7.1), quickly blotted dry, weighed and transferred to a respirometer (Oxygraph-2k; Oroboros Instruments, Innsbruck, Austria) in respiration solution at 37□°C. Oxygen concentration was maintained above 300□μM throughout the experiment to avoid limitations in oxygen supply. Background respiration was assessed before adding substrates and was subtracted from all subsequent values. Leak respiration (L) was assessed after the addition of sodium glutamate (10□mM), sodium malate (0.5□mM), and sodium pyruvate (5□mM). NADH (Complex I)-linked respiration was assessed after the addition of 5□mM ADP, and 10□μM cytochrome c to confirm the absence of outer-mitochondrial membrane damage. Maximal oxidative phosphorylation capacity, with simultaneous convergent electron input via NADH and FADH-linked pathways, was measured after the addition of 10□mM succinate. Maximum uncoupled respiration (E) was determined via titration of carbonylcyanide-4-trifluoro-methoxyphenylhydrazone (FCCP) in 0.5□µM steps until no further increase in oxygen consumption was observed. Succinate-linked respiration was measured after blocking mitochondrial complex I with 0.5□μM rotenone. Two measurements per sample were performed simultaneously, and the results were averaged. Respiration values were normalized to wet weight and expressed in pmol O_2_·s^−1^□mg^−1^. Samples that increased more than 10% from the addition of 5mM of ADP to 10 μM cytochrome c were excluded from analysis. E/L coupling efficiency was calculated as the difference between maximum uncoupled respiration and leak respiration divided by the maximum uncoupled respiration. The phosphorylation capacity through complex I (PN/PNS) and complex II (PS/PNS) were calculated as Complex I-linked respiration normalized to maximal oxidative phosphorylation capacity, and Complex II-linked respiration normalized to maximal uncoupled respiration, respectively.

### Immuno-histochemistry

To visualize muscle fiber type and cross-sectional area, immunohistochemistry was carried out on 10□µm thick sections from the vastus lateralis muscle biopsies. Details about dilutions and vendors can be found in Table 2.

**Table 2:** List of antibody vendors.

| **Parameter** | **Primary antibody, dilution, vendor** | **Catalog #/RRID** | **Secondary Antibody, dilution, vendor** | **Catalog #/RRID** |
| --- | --- | --- | --- | --- |
| **Muscle Fibre Type** | BA-D5, 1µg/mL, DSHB | DSHB Cat# BA-D5, RRID:AB_2235587 | Goat Anti-mouse IgG2B 555, 1:2000, Invitrogen | Thermo Fisher Scientific Cat# A-21147, RRID:AB_2535783 |
|  | 6H1, 5µg/mL, DSHB | DSHB Cat# 6H1, RRID:AB_1157897 | Goat Anti-mouse IgM 488, 1:2000, Invitrogen | Thermo Fisher Scientific Cat# A-21042, RRID:AB_2535711 |
|  | SC-71 1µg/mL, DSHB | DSHB Cat# SC-71, RRID:AB_2147165 | Goat Anti-mouse IgG1 647, 1:2000, Invitrogen | Thermo Fisher Scientific Cat# A-21240, RRID:AB_2535809 |
|  | WGA 350, 1:25, Invitrogen | W11263 |  |  |
| **Capillaries** | Ulex europaeus agglutinin I, 20µg/mL, Vector Laboratories | Vector Laboratories Cat# B-1065, RRID:AB_2336766 | Vectastain Elite ABC Kit PK-6100, Vector Laboratories ImmPACT™ AMEC Red Peroxidase Substrate, Vector Laboratories | Vector Laboratories Cat# PK-6100, RRID:AB_2336819<br>Vector Laboratories Cat# SK-4285, RRID:AB_2336519 |

### Fiber-type composition

Skeletal muscle fiber type quantification and size were assessed using immunofluorescence techniques using primary antibodies against myosin heavy chain (MHC) I (BA-D5), MHC IIa (SC-71), and MHC IIx (6H1)^6^. Firstly, sections were air-dried for 10□min and then blocked with 10% normal goat serum (NGS) for 60□min. Subsequently, slides were washed 3 times for 5□min each in 1x phosphate-buffered saline (PBS) and incubated with the primary antibodies for 60□min at room temperature. Slides were washed, and subsequently incubated with the secondary antibodies for 60□min in the dark at room temperature. Another washing step was performed and subsequently, the slides were incubated with Wheat Germ Agglutinin (WGA) for 30□minutes in the dark at room temperature. This was followed by a final washing step before the sections were mounted with coverslips using Vectashield Vibrance. Analysis was performed with ImageJ and a modified version of Sandia Matlab AnalysiS Hierarchy (SMASH) Toolbox (version 1.0) in Matlab (Version 2022a). Fiber type composition was expressed as the cross-sectional area occupied by each fiber type as a percentage of the cross-sectional area of all fibers. This allowed for quantification of fiber type specific fiber cross-sectional area.

### Succinate dehydrogenase activity

Sections were stained for SDH activity as described previously^6^. Immediately after cutting, sections were air-dried for 15□min and then immersed in a pre-heated (37□°C) solution containing sodium phosphate buffer (0.1□M, pH 7.6), sodium succinate (0.2□M), sodium azide (14□mM) and tetranitro blue tetrazolium (TNBT, 0.55□mM, Sigma-Aldrich) for 20□min in the dark, and the reaction was stopped by brief HCl (0.01□M) exposure before sections were washed and mounted with glycerin-gelatin. Sections were stored at 4□°C and imaged within ten days. The averaged SDH activity was obtained by outlining 40 individual skeletal muscle fibers per participant and absorbance was assessed using ImageJ (Version 1.54f, NIH, Bethesda, USA). Values were expressed as Δ*A*_660_ per µm tissue thickness per second of staining time (Δ*A*_660_□µm^−1^□s^−1^).

### Capillarization

Capillarization was assessed by staining for Ulex Europaeus Agglutinin 1 lectin (UEA-1)^6^. Sections were air-dried for 10□min and fixated in ice-cold acetone (−20□°C) for 15□min. Subsequently, slides were washed 3 times for 2□min in PBS and blocked with 1% bovine serum album for 30□min. Afterwards, slides were incubated with UEA-1 for 30□min at room temperature, followed by wash steps and incubation with Vectastain Elite ABC kit for 30□min at room temperature. After washing, incubation with Red Peroxidase substrate for 10□min at room temperature was performed, washed, and sections were mounted with glycerin-gelatin (heated at 37□°C). Capillary-to-fiber ratio and capillary density by capillaries per mm^2^ muscle tissue, and capillary cross sectional area were determined in ImageJ.

### Myoglobin Content

To assess intracellular myoglobin content, muscle biopsy sections were freeze-dried and fixed with paraformaldehyde (158127, Sigma-Aldrich) at 70-75°C for 60min using a vapor-fixation technique^73^. Slides were fixed with 2.5% glutaraldehyde (G5882, Sigma-Aldrich) buffer for 10min before a 60min incubation in O-tolidine-containing solution. The O-tolidine-containing solution was prepared in 62 ml 50 mM Tris-80 mM KCl buffer containing 25 mg ortho-tolidine (T8533, Sigma-Aldrich), dissolved in 2 ml 95% ethanol, and 1.43 ml 70% tertiary-butyl-hydroperoxide (458139, Sigma-Aldrich). Incubations were performed at 50 °C. Sections were washed and mounted with glycerin-gelatin. Ninety randomly selected fibers regardless of fiber type were analyzed using Fiji^74^. Values were expressed as arbitrary unit (AU).

### Image acquisition

Slides were dried overnight at 4□°C before imaging. Images for fiber type and capillarization analysis were taken at ×20 magnification with VS200 Research Slide Scanner (Olympus) using Slideview 5.0. SDH images were taken with a DMRB microscope (Leica, Wetzlar) using a CCD camera, calibrated gray filters, and an individual calibration curve at 660□nm. Myoglobin images were captured with ×20/0.4 objective using a CCD camera at 436±9nm absorbance. Approximately fifteen images were taken per sample, with 5 images per slide-section. All images were manually evaluated to exclude those containing out-of-focus areas, artifacts, and large areas of connective tissue.

### Statistical analysis

Distributions were assessed with histograms and Shapiro–Wilk tests. Parametric quantitative variables are presented as means□±□standard deviation (SD), and nonparametric quantitative variables are presented as median and interquartile ranges (IQR; 25th and 75th percentiles). Jitter plots are plotted with means represented as single bars. All individual points represent a participant.

Non-normally distributed data were transformed using Box-Cox transformation in order to obtain a normal distribution for statistical analysis. If data remained non-normally distributed following Box-Cox transformations, group differences were compared using a Kruksal-Wallis test, with pairwise Wilcoxon signed-rank test post-hoc where applicable. Continuous parametric data were analyzed using a paired *t*-test for bed rest data or analysis of variance, with Tukey HSD post-hoc testing, where appropriate. Continuous nonparametric data were analyzed using the Mann–Whitney *U* test, Kruskal– Wallis *H* test, or pairwise Wilcoxon test with Benjamini–Hochberg correction where appropriate.

Correlations were analyzed using the Pearson correlation coefficient for linear relationships. Correlation coefficients were compared using the R package *cocor*. All data were analyzed using R studio built under R version 4.0.3 (R Core Team 2013, Vienna, Austria).

### Reuse of previously published data

Some results from bed rest participants were reanalyzed from previously published data ^22,75^. Biopsies were restained and analyzed for capillarization, fiber type, and myoglobin content to account for differences in staining protocols and analysis procedures. A part of the results from patients with long COVID and 21 healthy controls were previously published^6^.

## Data Availability

The source data file is available upon request from the corresponding author.

## Author Contributions

Study design and concept: BTC, BA, RPG, HD, MvV, RCIW

Data acquisition: BTC, AS, BA, RPG, JYH, ME, WN, PWH, FWB, JJP, PvA,

Data analysis: BTC, AS, JH, RPG, EAB, ME, TK, LV, JV, LG

Writing manuscript: BTC, AS, BA, JYH, ME, WN, PWH, FWB, JJP, PvA, RPG, HD, MvV, RCIW

## Acknowledgements

We thank Ellen A. Breedveld, Esmee van den Berg, Tom Kerkhoff, Augustijn M Klarenbeek, Jessie Hulscher, and Noor Bijvang for their help during the data collection of the long COVID and ME/CFS patients, and Bergita Ganse, Alessandra Bosutti, Edwin Mulder and Jörn Rittweger for their help in the AGBRESA bed rest study. The authors want to thank our patient representatives for insightful discussions.

