## Supplemental file for "Skeletal muscle properties in long COVID and ME/CFS differ from those induced by bed rest"

**Supplemental Table 1**: Sub-cohort characteristics, data presented as median (IQR). All data remained non-normally distributed following Box-Cox transformations. Group differences were compared using a Kruksal-Wallis test, with pairwise Wilcoxon signed-rank test post-hoc where applicable. ME/CFS: myalgic encephalomyelitis/chronic fatigue syndrome.

|  | Bed rest | Healthy Control | Bed rest | Long COVID | ME/CFS |
| --- | --- | --- | --- | --- | --- |
|  | **Pre** |  | **Post** |  |  |
| Participants (# Females) | 17  (7) | 17  (7) | 13  (7) | 13  (7) | 13  (7) |
| Age  (years) | 34  (29-43) | 34  (29-42) | 29  (27-40) | 31  (28-42) | 32  (28-42) |
| Height  (cm) | 175  (172-179)  * | 180  (176-186) | 174  (162-175) | 177  (168-191) | 175  (172-186) |
| Weight  (kg) | 76  (69-80) | 74  (63-86) | 70  (66-76) | 82  (71-90) | 84  (75- 86) |
| Symptom Duration (days) | n/a | n/a | n/a | 500  (408-599) ††† | 3793  (2463-4613) |
| Daily Steps (steps*day^-1^) | ND | 7011  (4770-8316) | 0 | 3970  (2794-5030) | 3827  (2722-4901) |
| *: *vs.* healthy control; †: *vs.* ME/CFS; §: *vs*. Bed rest  *: p<0.05; **: p<0.01; ***: p<0.001 | | | | | |

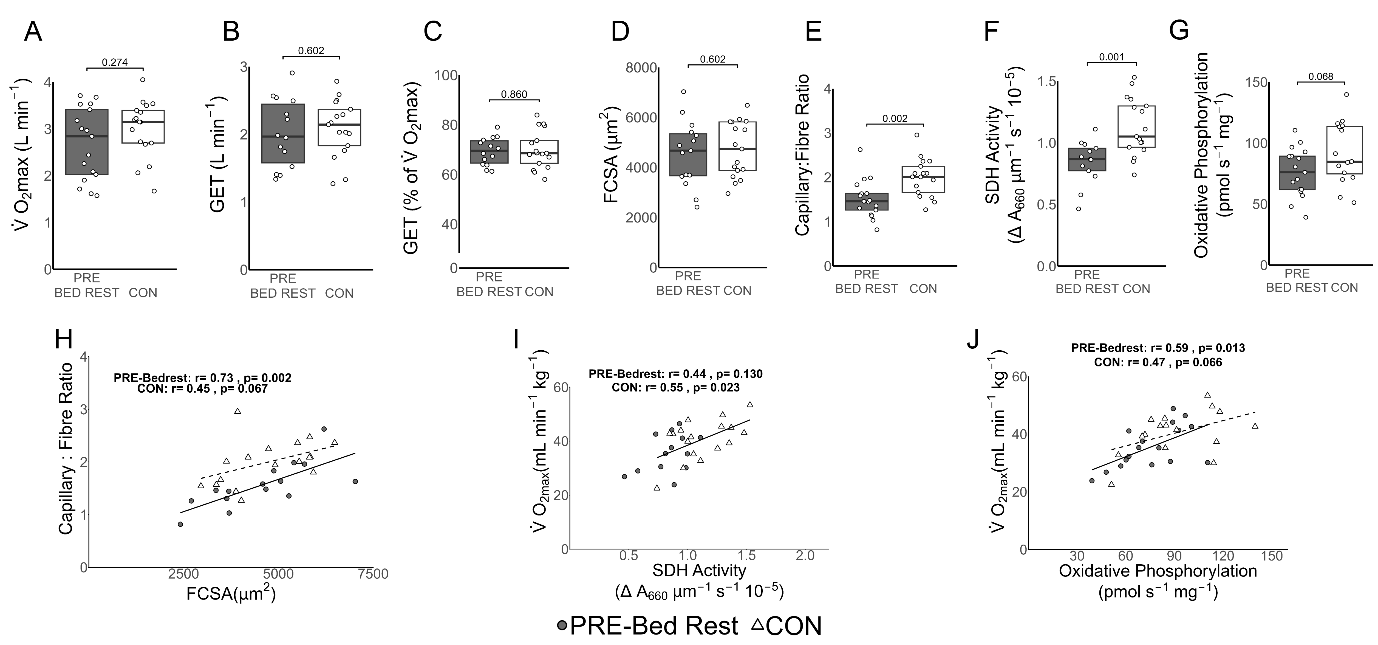

**Supplemental Figure 1**: A sub-analysis was conducted on age- and sex-matched individuals between healthy individuals pre-bed rest (n=17) and healthy individuals matched to patients with long COVID and ME/CFS (n=17). There were no differences in $\dot{V}O_{2max}$ (**A**), in gas exchange threshold (GET) (**B**), the occurrence of the gas exchange threshold relative to $\dot{V}O_{2max}$ (**C**) or fiber cross sectional area (FCSA, **D**) between groups. Participants before bed rest had a lower capillary-to-fiber ratio (**E**), lower succinate dehydrogenase (SDH) activity (**F**), and oxidative phosphorylation capacity (**G**) than patient-matched healthy controls. Both groups had, or tended to have a significant relation between FCSA and capillary-to-fiber ratio (**H**). Only patient-matched healthy controls had a significant relation between $\dot{V}O_{2max}$ and SDH activity (**I**). Both groups had, or tended to have, a significant association between $\dot{V}O_{2max}$ and oxidative phosphorylation capacity (**J**). Data for $\dot{V}O_{2max}$, FCSA, and SDH activity remained non-normally distributed after Box-Cox transformations. Parametric comparisons were assessed using non-paired t-tests. Non-parametric comparisons were assessed using Wilcoxon signed rank tests. Solid lines represent significant correlations, dashed lines represent correlations with *P*<0.10.

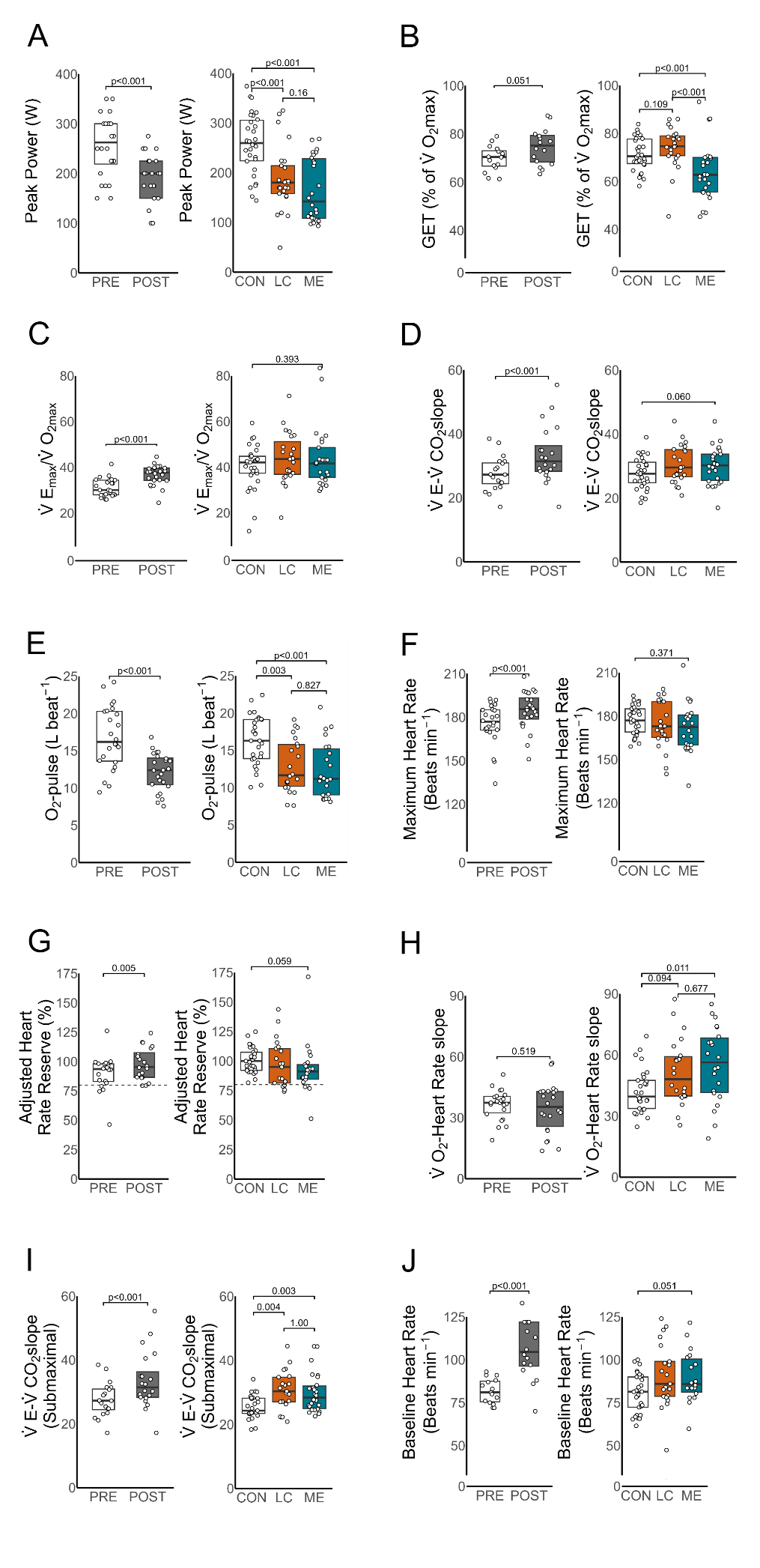

**Supplementary Figure 2**: Cardiopulmonary exercise test (CPET) data from pre- and post-bed rest, healthy controls, patients with long COVID and patients with ME/CFS. Peak power was reduced following bed rest (**A**), and was also lower in patients with long COVID and ME/CFS. Following bed rest, $\dot{V}E_{max}/\dot{V}CO_{2max}$ (**B**), $\dot{V}E_{max}/\dot{V}O_{2max}$ (**C**), and $\dot{V}E/\dot{V}CO_{2}$ slope (**D**) all increased, whereas patients with long COVID and ME/CFS were not significantly different from healthy controls. O_2_-pulse was significantly lower following bed rest (**E**), and lower in patients with long COVID and patients with ME/CFS compared to healthy controls. Maximal heart rate was increased following bed rest, while patients with long COVID and patients with ME/CFS had similar maximal heart rates to healthy controls (**F**). Following bed rest, adjusted heart rate reserve (AHRR) was significantly higher, but patients with ME/CFS had the tendency have a lower AHRR compared to healthy controls (**G**). the $\dot{V}O_{2}$-heart rate slope was not altered following bed rest, while both patients groups had, or tended to have, a higher $\dot{V}O_{2}$-heart rate slope compared to healthy controls (**H**). After bed rest, the gas exchange threshold (GET) tended to occur at a higher percentage of $\dot{V}O_{2max}$, whereas the GET occurred at a lower percentage of $\dot{V}O_{2max}$ in patients with ME/CFS compared to both healthy controls and patients with long COVID (**I**). Submaximal $\dot{V}E/\dot{V}CO_{2}$ slope was increased following bed rest, and also increased in both patients groups compared to controls (**J**). Baseline heart rates were elevated following bed rest (**K**), and tended to be higher in patients with long COVID and patients with ME/CFS than healthy matched controls. Comparisons between pre- and post- bed rest were assessed using paired t-tests for parametric data and Mann–Whitney *U* test for non-parametric data. Comparisons between patients with long COVID and patients with ME/CFS and healthy controls were assessed using ANOVA, with Tukey HSD post-hoc testing for parametric data or Kruskal–Wallis *H* test, with pairwise Kruskal–Wallis tests with Benjamini–Hochberg correction post-hoc for non-parametric data.

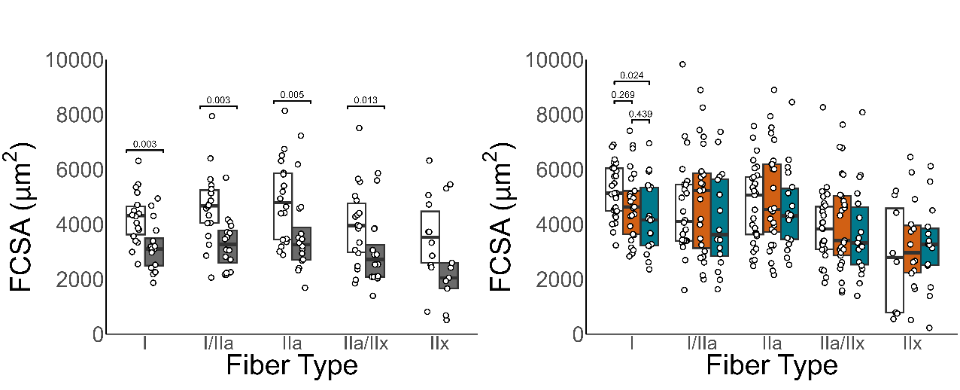

**Supplementary Figure 3**: Average fiber cross-sectional area (FCSA) for each individual fiber type was reduced from pre- to post-bed rest, with the exception of the few pure type IIx fibers. Type I fiber FCSA was significantly lower in patients with ME/CFS (green) compared to healthy controls, while patients with long COVID (orange) were not significantly different from healthy controls. After Box-Cox transformations, data for FCSA of type I, type I/IIa and IIa fibers were normally distributed, while data for FCSA of type IIa/IIx and IIx fibers remained non-normally distributed. Comparisons between pre- and post- bed rest were assessed using paired t-tests for parametric data and Mann–Whitney *U* test for non-parametric data. Comparisons between patients with long COVID and patients with ME/CFS and healthy controls were assessed using ANOVA, with Tukey HSD post-hoc testing for parametric data or Kruskal–Wallis *H* test, with pairwise Wilcoxon tests with Benjamini–Hochberg correction post-hoc for non-parametric data.

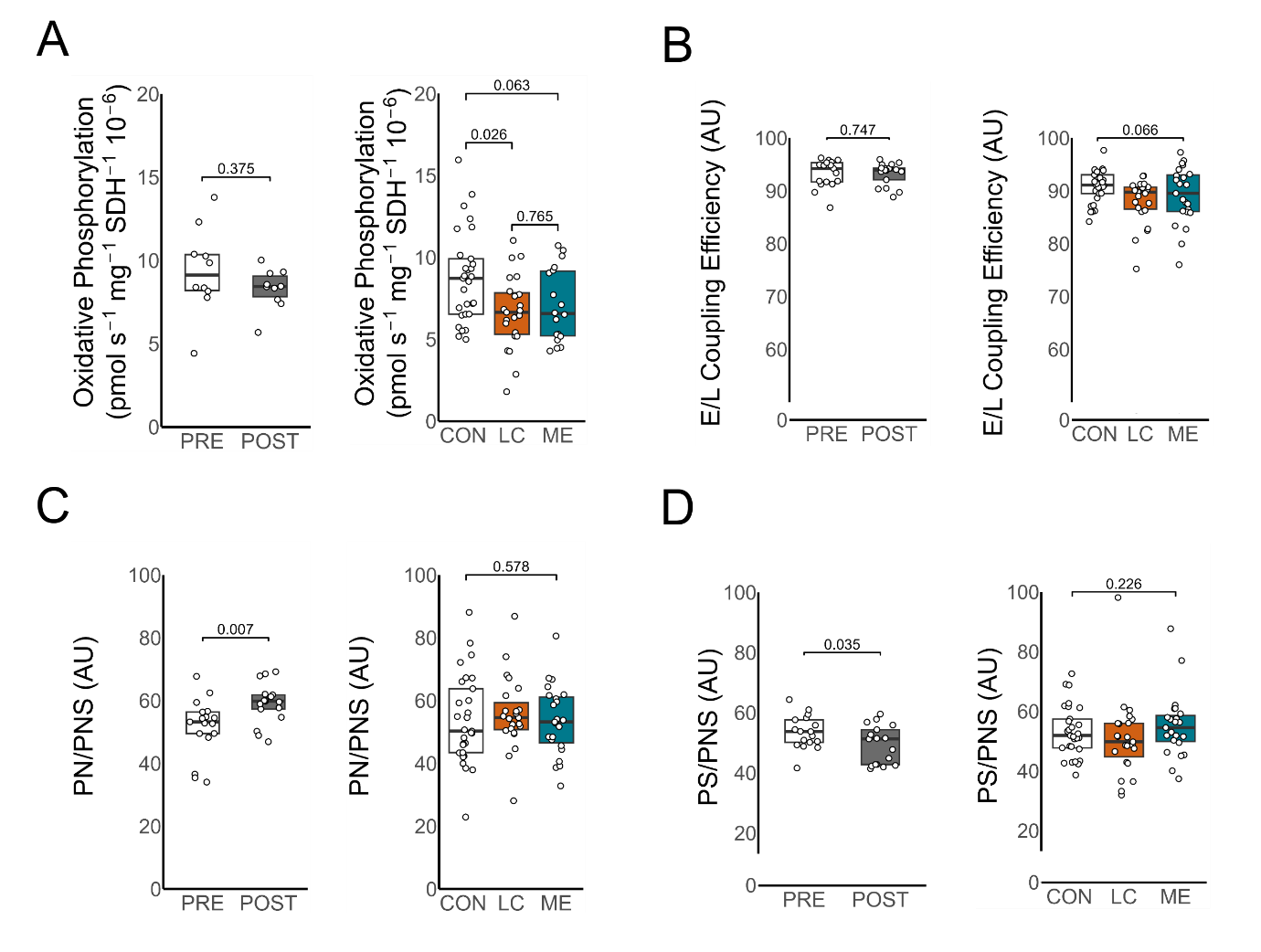

**Supplementary Figure 4**: Oxidative phosphorylation capacity was normalized to succinate dehydrogenase (SDH) activity to provide a marker for intrinsic mitochondrial respiration. Bed rest did not alter the normalized oxidative phosphorylation capacity, while long COVID (LC) and ME/CFS patients displayed lower capacities than healthy controls (**A**). Bed rest did not alter biochemical coupling (uncoupled/leak (E-L) coupling efficiency, while patients tended to have lower biochemical coupling compared to healthy controls (**B**). Bed rest increased the NADH-linked respiration flux control ratio (PN/PNS; **C**) and decreased the succinate-linked respiration flux control ratio (PS/PNS; **D**), whereas patients were not different from healthy controls. After Box-Cox transformations, all data remained non-normally distributed. Data from bed rest participants were compared using a paired Wilcoxon Signed Rank test. Data from patients with long COVID and ME/CFS and healthy controls were compared using a Kruksall-Wallis *H* test, and pairwise Wilcoxon Signed rank test post-hoc where applicable.

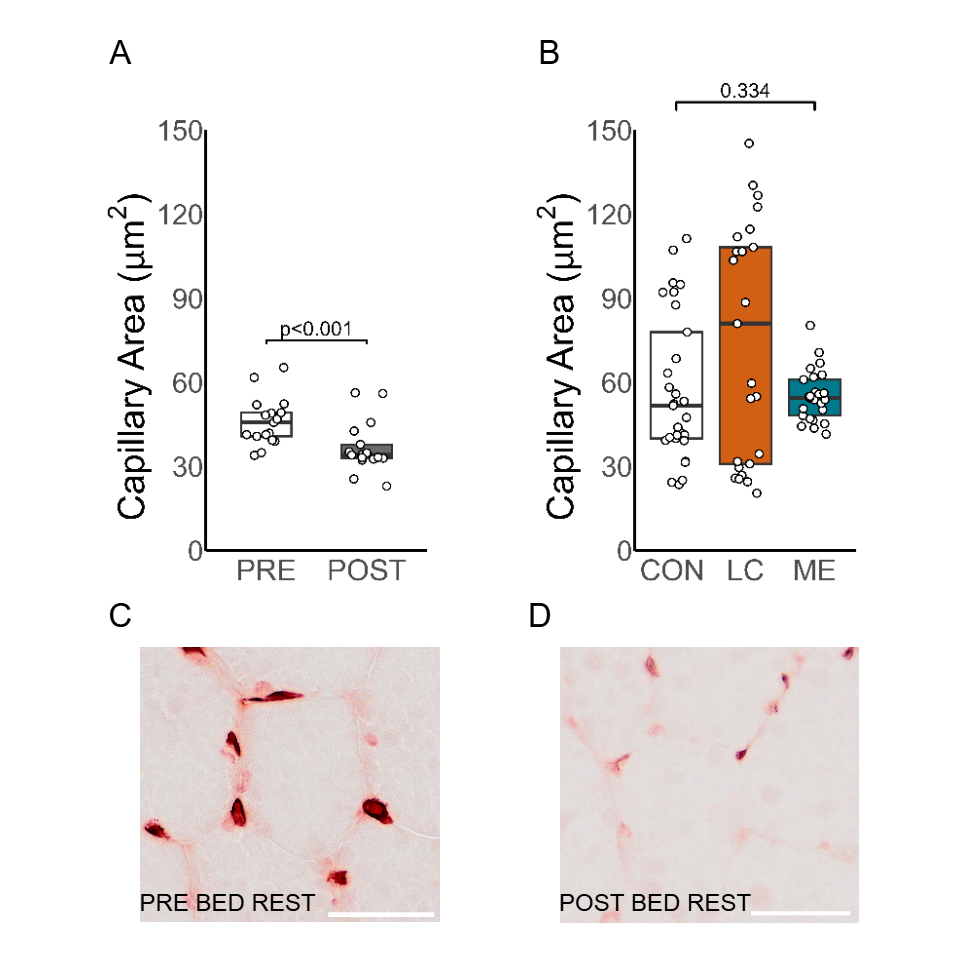

**Supplementary Figure 5**: Average capillary area decreased significantly following bed rest (**A**). Capillary area was not different between healthy controls and patients with ME/CFS or long COVID (LC, **B**). Typical examples of capillary staining in bed rest are shown in **C**. After Box-Cox transformations, capillary area data from bed rest participants was normally distributed, but data from patients with ME/CFS and long COVID and healthy controls remained non-normally distributed. Data from patients with ME/CFS and long COVID and healthy controls were compared using a Kruksall-Wallis *H* test.

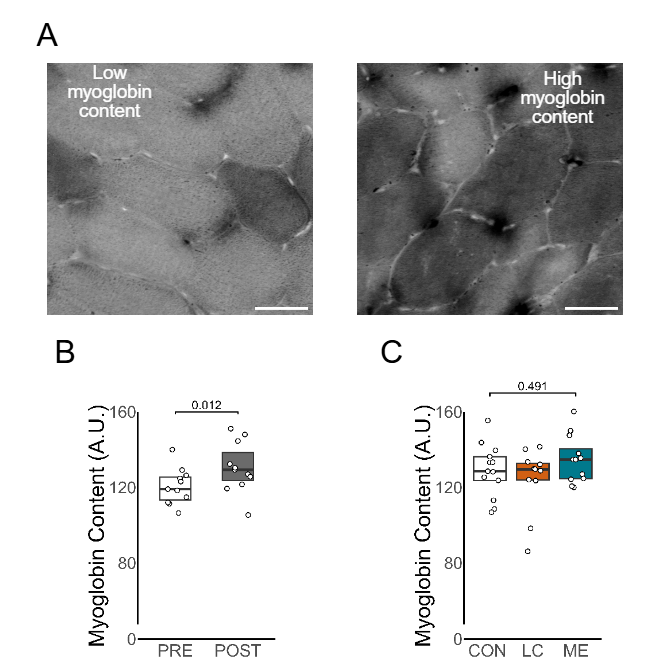

**Supplementary Figure 6**: Typical examples of low myoglobin content and high myoglobin content are displayed (**A**). Myoglobin content was measured in a subset of participants. Following bed rest myoglobin content was significantly elevated (**B**; n=11), while no differences were found between healthy controls **(C**; n=13), patients with long COVID (LC, n=11), and patients with ME/CFS (n=12). Scale bar represents 50 µm. After Box-Cox transformations, myoglobin data from bed rest participants was normally distributed, but data from patients with ME/CFS and long COVID and healthy controls remained non-normally distributed. Data from bed rest participants were compared using a paired t-test. Data from patients with ME/CFS and long COVID and healthy controls were compared using a Kruksall-Wallis *H* test.

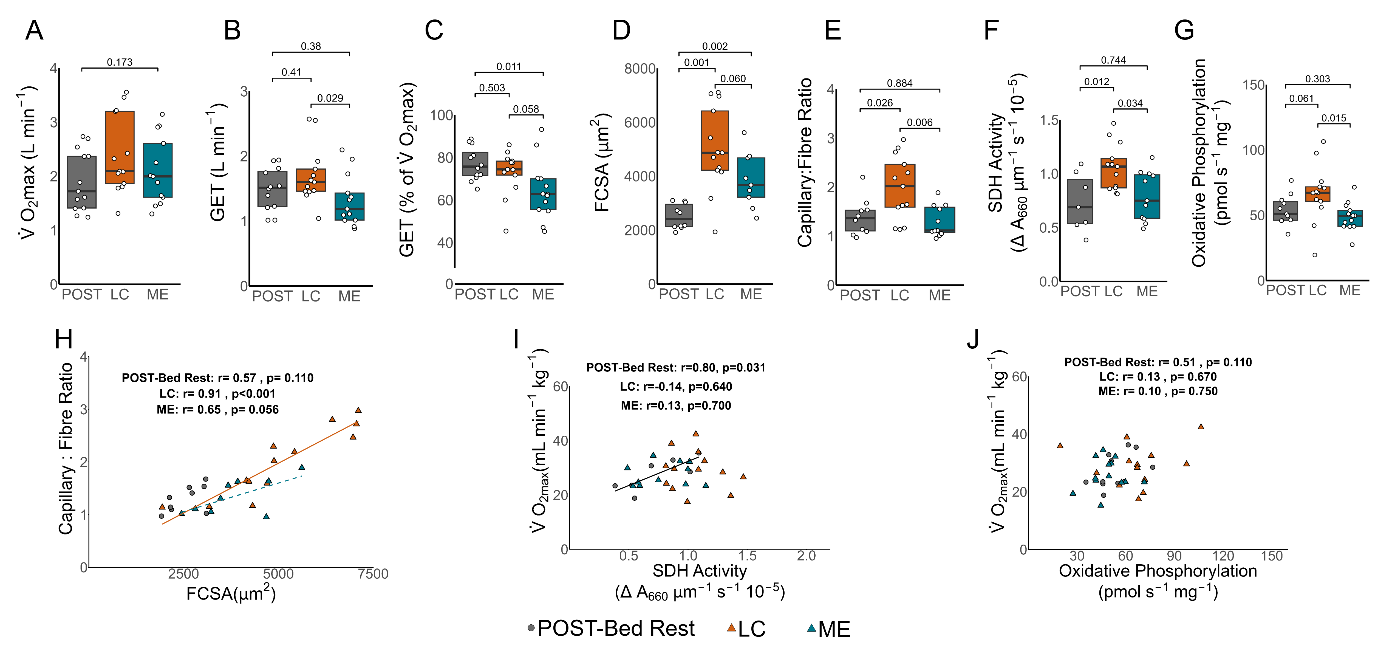

**Supplemental Figure 7**: A sub-analysis was conducted on age- and sex-matched individuals between post-bed rest (n=13), patients with long COVID (LC, n=13) and patients with ME/CFS (n=13). There were no differences in $\dot{V}O_{2max}$ between groups (**A**), however the gas exchange threshold was lower in patients with ME/CFS than patients with long COVID (**B**). The gas exchange threshold relative to $\dot{V}O_{2max}$ was also earlier in patients with ME/CFS compared to post-bed rest and patients with long COVID (**C**). Skeletal muscle fiber cross-sectional area (FCSA) was lower after bed rest compared to either ME/CFS or long COVID, while there was a trend for patients with ME/CFS to present with smaller fibers than patients with long COVID (**D**). Capillary-to-fiber ratio was higher in long COVID than either post-bed rest or patients with ME/CFS (**E**). Similarly, succinate dehydrogenase (SDH) activity (**F**) and oxidative phosphorylation capacities (**G**) were higher in patients with long COVID than either post-bed rest or patients with ME/CFS. All groups had, or tended to have, a significant relation between FCSA and capillary-to-fiber ratio (**H**). There was a (weak) correlation between $\dot{V}O_{2max}$ and either SDH activity (**I**) or oxidative phosphorylation capacity (**J**) in participants after bed rest, but not in either patient group. Data from $\dot{V}O_{2max}$, gas exchange threshold relative to $\dot{V}O_{2max}$, FCSA, and oxidative phosphorylation remained non-normally distributed after Box-Cox transformations. Parametric comparisons were assessed using ANOVA, with Tukey HSD post-hoc testing. Non-parametric comparisons were assessed using Kruskal–Wallis *H* test, with pairwise Wilcoxon tests with Benjamini–Hochberg correction post-hoc. Solid lines represent significant correlations, dashed lines represent correlations with *P*<0.10.
